# Associations of high-density lipoprotein cholesterol concentration and particle count with mortality: a prospective study of 50,000 men and 100,000 women

**DOI:** 10.64898/2026.09.24.26363978

**Authors:** Gary O’Donovan, Shoaib Afzal, Fanny Petermann-Rocha, Evelia Apolinar-Jiménez, Catalina Medina, Pablo Kuri-Morales, Roberto Tapia-Conyer, Jesús Alegre-Díaz

**Affiliations:** Facultad de Medicina, Universidad de los Andes, Bogotá, Colombia; Department of Clinical Biochemistry, Copenhagen University Hospital – Herlev and Gentofte, Copenhagen, Denmark; Department of Clinical Medicine, Faculty of Health and Medical Sciences, University of Copenhagen, Copenhagen, Denmark; Centro de Investigación Biomédica, Facultad de Medicina, Universidad Diego Portales, Santiago, Chile; Departamento de Investigación, Hospital Regional de Alta Especialidad del Bajío adscrito a Servicios de Salud del Instituto Mexicano del Seguro Social para el Bienestar (IMSS-Bienestar), León, México; Centro de Investigación en Nutrición y Salud, Instituto Nacional de Salud Pública, Cuernavaca, Morelos, México; Facultad de Medicina, Universidad Nacional Autónoma de México (UNAM), Ciudad de México, México; Instituto Tecnológico y de Estudios Superiores de Monterrey, Monterrey 64849, México

**Author notes:** Correspondence: Dr. Gary O’Donovan, Facultad de Medicina, Universidad de los Andes, Carrera 1, 18A – 12, Bogotá, Colombia.

## Abstract

**Background and Objectives:** More research is required to determine the associations of high-density lipoprotein (HDL) cholesterol concentration and particle count with mortality. The objectives of this analysis were to investigate whether there were non-linear associations of HDL concentration with all-cause mortality, whether cardiovascular disease (CVD) deaths, cancer deaths or other deaths drove these associations, and whether small, medium, or large HDL particle count was associated with mortality.

**Methods and Results:** The sample included 47,123 men and 97,589 women from the Mexico City Prospective Study. Models were adjusted for confounders and competing risks. There were 11,264 deaths in men and 15,486 in women. There was a J-shaped association of HDL concentration with all-cause mortality in men driven by other deaths. For example, compared with the >40^th^-60^th^ percentile, the hazard ratio (95% confidence interval) for other mortality was 1.27 (1.11, 1.44) in the >95^th^-99^th^ percentile and 1.82 (1.45, 2.28) in the >99^th^-100^th^ percentile. There was a U-shaped association in women driven by CVD deaths at lower concentrations and other deaths at higher concentrations: the hazard ratio for CVD mortality was 1.23 (1.05, 1.43) in the two lowest percentiles groups and the hazard ratio for other mortality was 1.33 (1.18, 1.50) in the two highest percentile groups. Small HDL particle count was associated with reduced risk of all-cause, CVD, and other mortality in men and women. For example, compared with the bottom quartile, the hazard ratio for all-cause mortality was 0.79 (0.73, 0.86) in the top quartile in men and 0.77 (0.72, 0.82) in the top quartile in women.

**Conclusions:** There is a J-shaped association of HDL concentration with mortality in men and a U-shaped association in women. More research is required to confirm the novel finding that small HDL particle count reduces the risk of all-cause, CVD, and other mortality in men and women.

## Introduction

High-density lipoprotein (HDL) cholesterol is commonly known as ‘good cholesterol’ because higher concentrations are thought to reduce the risk of cardiovascular disease (CVD) [1–3]. Early studies tended to show that ‘high’ HDL cholesterol concentrations were associated with lower risk of CVD morbidity and CVD mortality than ‘medium’ concentrations or ‘low’ concentrations [4]. However, the earlier studies were relatively small and were not designed to distinguish between high concentrations and extremely high concentrations and between low concentrations and extremely low concentrations [4]. More recent studies are relatively large and tend to show that the associations of HDL cholesterol concentration with all-cause mortality and CVD mortality are U-shaped, with both extremely low concentrations and extremely high concentrations associated with greater risks [5]. HDL cholesterol is also known as good cholesterol because it is thought to be involved in reverse cholesterol transport [1–3]. However, it has been suggested that HDL particles may become dysfunctional and may not facilitate the removal of cholesterol from macrophages in arterial walls [5–7]. Small functional particles may become large dysfunctional particles [6, 7], and there is some evidence that small HDL particle count is associated with lower risk of cardiovascular events [8].

More research is required to clarify the associations of HDL cholesterol concentration with mortality because even some of the largest studies were not stratified by sex [9], some were not adjusted for triglycerides and other major confounders [10, 11], and some did not adequately address the problem of reverse causation by excluding participants who died during the early years of follow-up [12–14]. More research is also required to determine whether CVD deaths, cancer deaths, or other deaths drive the apparent U-shaped associations of HDL cholesterol concentration with mortality because some studies did not treat different causes of death as competing risks [10, 11, 15, 16]. Finally, while there is some evidence that small HDL particle count is associated with lower risk of cardiovascular events [8], there is little evidence about the associations of HDL particle count with mortality [16]. This study had three objectives in light of the available evidence. The first objective was to investigate whether there were non-linear associations of HDL cholesterol concentration with all-cause mortality. The second objective was to investigate whether CVD deaths, cancer deaths, or other deaths were driving any non-linear associations of HDL cholesterol concentration with all-cause mortality. The third objective was to investigate the associations of small, medium, and large HDL particle counts with mortality.

## Methods

### Participants

Participants were from the Mexico City Prospective Study, which is described in great detail in the study profile [17] and the data showcase [18]. Men and women were recruited from the district of Coyoacán and the less affluent district of Iztapalapa. The baseline survey was from 1998 to 2005 and the resulting sample was deemed to be representative of the population aged 35 years or older by virtue of the large sample size of more than 150,000 adults and the high response rate of more than 90% of eligible households [17]. The study was approved by the Mexican Ministry of Health, the Mexican National Council of Science and Technology (approval number 0595 P-M), and the Central Oxford Research Ethics Committee (C99.260). Trained nurses collected data in the participants’ homes and all participants provided informed consent.

### Exposures

The exposures were HDL cholesterol concentration and HDL particle size. Blood samples were taken, stored temporarily at 4-10 °C, and stored overnight at 4 °C. Plasma samples were then held in ultra-low temperature freezers in a central laboratory in Mexico before being shipped on dry ice to Europe [17]. Around 80% of the samples were analysed in a laboratory in Finland and 20% in a laboratory in the UK. The Nightingale Health nuclear magnetic resonance (NMR) platform was used in both laboratories [19]; And, biomarker levels were recalibrated to remove inter-spectrometer variation [18]. The median variance before recalibration was 3.2% for all 107 ‘non-derived’ NMR biomarkers, and was 6.0% for small HDL particles, 4.6% for medium HDL particles, and 2.9% for large HDL particles. Participants were not asked to fast before giving blood.

### Outcomes

The outcomes were all-cause mortality, CVD mortality, cancer mortality, and ‘other mortality’ (i.e. all non-CVD and non-cancer deaths). Mortality was tracked to 30^th^ September 2022 through probabilistic linkage to the national death register based on the participant’s name, age and sex. The registration of deaths is thorough in Mexico, with almost all deaths certified medically and with few deaths attributed to unknown causes [20]. Diseases listed on the death certificates were coded according to the International Classification of Diseases, 10th Revision [18, 21].

### Potential confounders

Sex is probably a major confounder because of the considerable biological and sociological differences between men and women. For example, average HDL cholesterol concentrations may be different in men and women [22–24], including adults of Latin American origin [24–26]. CVD risk is also greater in men than in women at any given age [27]. And, men and women often have different gender roles that affect their physical activity levels and other lifestyle choices [28–31]. Other potential confounders that may influence the associations of HDL with mortality include age, triglycerides, low-density lipoprotein (LDL) cholesterol, lipid medication, blood pressure, diabetes, socioeconomic status, smoking, alcohol, diet, body mass index, and leisure time physical activity [5, 7]. The nurse asked the participant how old they were and what sex they were. Then the nurse asked the participant about their highest level of education, their main occupation, their monthly income, and whether they took part in any sport or exercise. Finally, the nurse asked about smoking habits, drinking habits, medical history, fruit and vegetable intake, and medication use [17, 18]. At the end of the interview, the nurse measured blood pressure on three occasions and weight and height in light clothing and without shoes. Finally, the nurse took a blood sample [17, 18]. The same NMR platform was used to assess HDL cholesterol concentration, HDL particle size, triglyceride concentration and LDL cholesterol concentration [19].

### Statistical analyses

Analyses were performed using Stata MP Version 18 for Mac (StataCorp, Texas, USA). Probability values of less than 5% were considered statistically significant (i.e. P<0.05). Analyses were conducted in men and women separately because stratification is probably the only way of eliminating the confounding effects of sex [32]. Data from the first two years of follow-up were not used to minimise the possibility of reverse causation. The present analysis did not include participants with extreme values for systolic blood pressure of >250 mm Hg or extreme values for body mass index of <15 or >60 kg/m^2^ [33, 34]. Histograms of HDL cholesterol concentrations and HDL particle sizes were created and all graphs were normally distributed or were bell-shaped and only slightly skewed; therefore, the data were not transformed.

The first objective of the present study was to investigate whether there were non-linear associations of HDL cholesterol concentration with all-cause mortality. To that end, restricted cubic splines were used to analyse the shape of the associations of HDL cholesterol concentration as a continuous variable with all-cause mortality. Akaike information criteria were used to evaluate the goodness of fit of splines with three to five knots.[35] The spline with the fewest knots was chosen if the Akaike information criteria were within two of each other in order to achieve a balance between best fit and overfitting [35]. The splines were incorporated in Cox proportional hazards models that were adjusted for the potential confounders of age, triglyceride concentration, LDL concentration, lipid medication, systolic blood pressure, diabetes, education, income, area of residence, smoking, alcohol, fruit and vegetable intake, body mass index, and leisure time physical activity at baseline. Age, triglyceride concentration, LDL cholesterol concentration, systolic blood pressure, and body mass index were treated as continuous variables and all other variables were treated as categorical.

The second objective was to investigate whether CVD deaths, cancer deaths, or other deaths were driving any non-linear associations of HDL cholesterol concentration with all-cause mortality. Cox proportional hazards models were used to analyse associations of HDL cholesterol concentration as a categorical variable with cause-specific mortality. HDL cholesterol concentration was expressed in nine percentile groups [15], and the reference group was the percentile group containing the concentration associated with the lowest risk of all-cause mortality as determined in the restricted cubic spline analyses. For CVD mortality, all non-CVD deaths were treated as competing risks [36]. For cancer mortality, all non-cancer deaths were treated as competing risks [36]. For other mortality, all CVD deaths and cancer deaths were treated as competing risks [36]. There were relatively few deaths in the lowest and highest percentile groups; therefore, to increase statistical power, we also investigated the risk of death in the two lowest percentile groups combined and in the two highest percentile groups combined. All the models were adjusted for the potential confounders. The proportional hazards assumption was checked graphically for the discrete percentile groups and no violations were observed.

The third objective was to investigate the associations of small, medium, and large HDL particle counts with mortality. Cox proportional hazards models were used to analyse the risk of death in the bottom quartile of particle count compared with the top quartile [8]. Again, CVD deaths, cancer deaths, and other deaths were treated as competing events [36]. Similarly, all the models were adjusted for the potential confounders. The proportional hazards assumption was checked graphically for the discrete quartile groups and no violations were observed.

## Results

Supplementary Figure S1 in the online supplement shows the flow of participants. The present analysis included data from 47,123 of 52,579 (90%) male cohort members, after excluding 1,130 men with less than two years of follow-up, 2,038 men lost to follow-up, and 2,288 men with any missing or extreme values for the exposures or confounders. The present analysis also included data from 97,589 of 106,938 (91%) female cohort members, after excluding 1,347 women with less than two years of follow-up, 3,088 women lost to follow-up, and 4,914 with any missing or extreme values for the exposures or confounders. Table 1 shows participants’ characteristics at baseline. In men, the average age was 53 years, the average HDL cholesterol concentration was 0.93 mmol/L, and the average triglycerides concentration was 1.64 mmol/L. It is notable that more than 60% of men were in the highest income tertile, that nearly 50% were smokers, and that around 30% took part in sport and exercise in their leisure time. In women, the average age was 53 years, the average HDL cholesterol concentration was 1.03 mmol/L, and the average triglycerides concentration was 1.53 mmol/L. It is notable that more than 60% of women were in the lowest income tertile, only 23% were smokers, and only 19% took part in sport and exercise. Less than one per cent of men and women reported taking lipid medication.

**Table 1.**
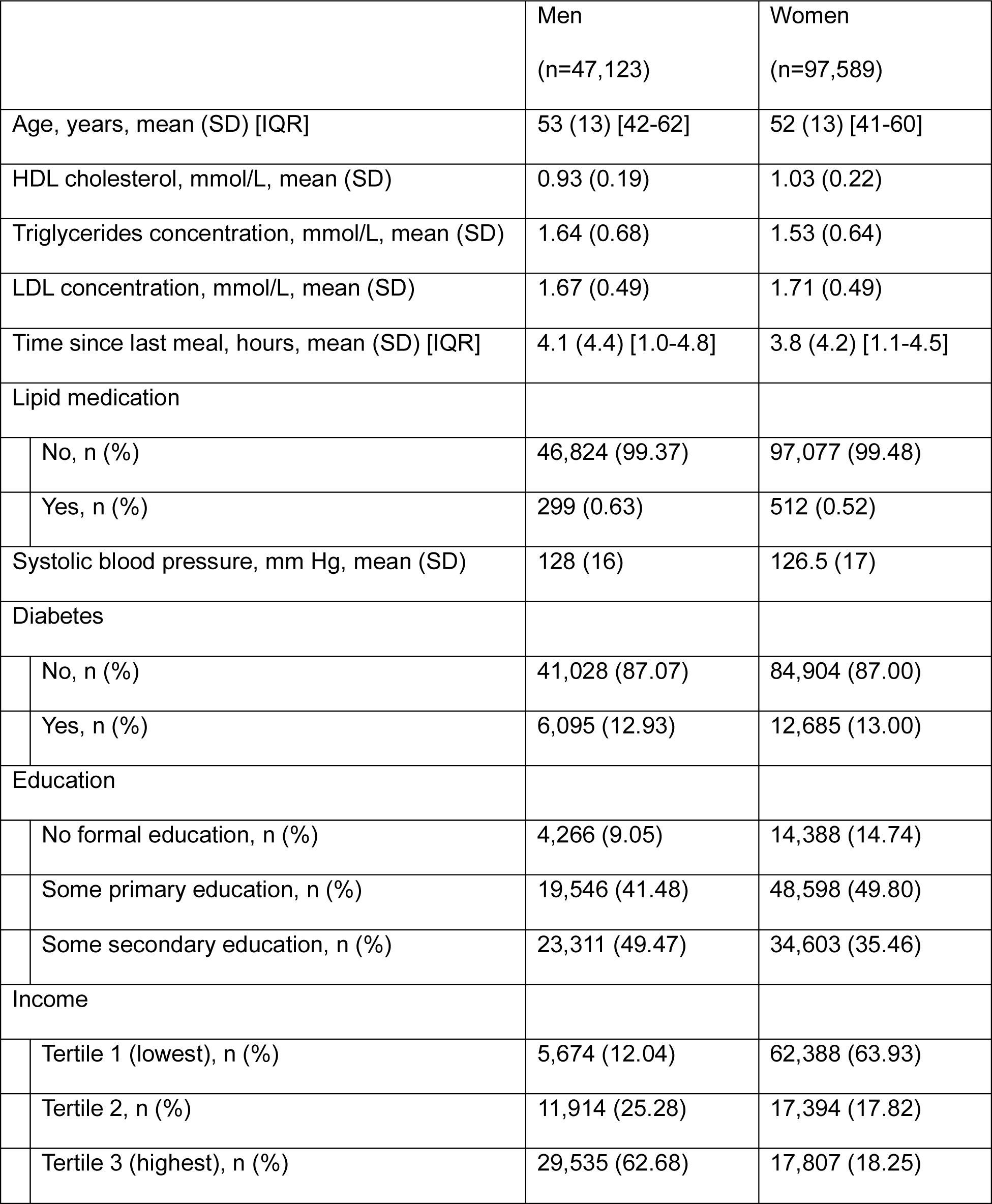
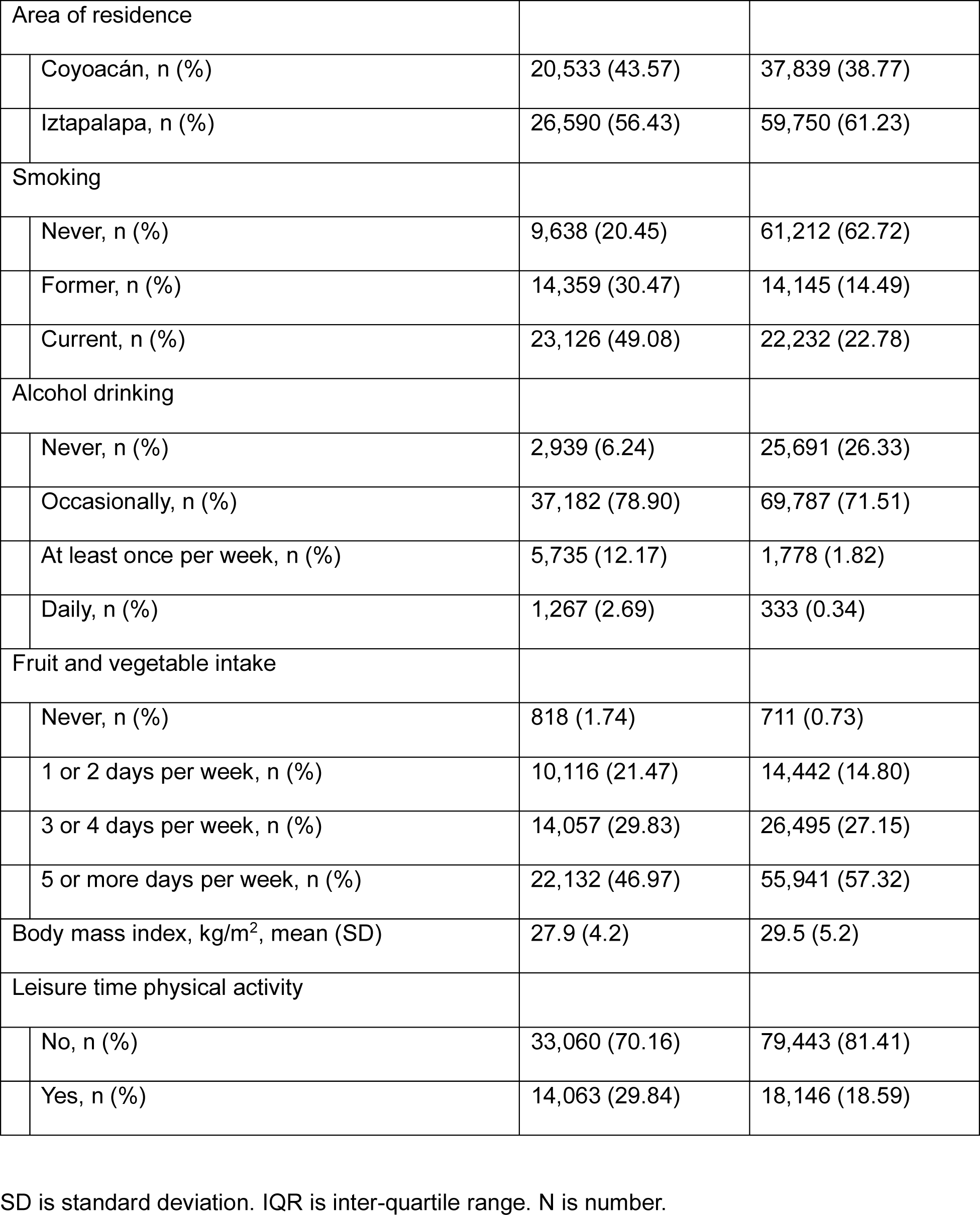
Participants’ characteristics at baseline according to sex.

|  | Men<br>(n=47,123) | Women<br>(n=97,589) |
| --- | --- | --- |
| Age, years, mean (SD) [IQR] | 53 (13) [42-62] | 52 (13) [41-60] |
| HDL cholesterol, mmol/L, mean (SD) | 0.93 (0.19) | 1.03 (0.22) |
| Triglycerides concentration, mmol/L, mean (SD) | 1.64 (0.68) | 1.53 (0.64) |
| LDL concentration, mmol/L, mean (SD) | 1.67 (0.49) | 1.71 (0.49) |
| Time since last meal, hours, mean (SD) [IQR] | 4.1 (4.4) [1.0-4.8] | 3.8 (4.2) [1.1-4.5] |
| Lipid medication |  |  |
| No, n (%) | 46,824 (99.37) | 97,077 (99.48) |
| Yes, n (%) | 299 (0.63) | 512 (0.52) |
| Systolic blood pressure, mm Hg, mean (SD) | 128 (16) | 126.5 (17) |
| Diabetes |  |  |
| No, n (%) | 41,028 (87.07) | 84,904 (87.00) |
| Yes, n (%) | 6,095 (12.93) | 12,685 (13.00) |
| Education |  |  |
| No formal education, n (%) | 4,266 (9.05) | 14,388 (14.74) |
| Some primary education, n (%) | 19,546 (41.48) | 48,598 (49.80) |
| Some secondary education, n (%) | 23,311 (49.47) | 34,603 (35.46) |
| Income |  |  |
| Tertile 1 (lowest), n (%) | 5,674 (12.04) | 62,388 (63.93) |
| Tertile 2, n (%) | 11,914 (25.28) | 17,394 (17.82) |
| Tertile 3 (highest), n (%) | 29,535 (62.68) | 17,807 (18.25) |

|  |  |  |  |
| --- | --- | --- | --- |
| Area of residence |  |  |  |
|  | Coyoacán, n (%) | 20,533 (43.57) | 37,839 (38.77) |
|  | Iztapalapa, n (%) | 26,590 (56.43) | 59,750 (61.23) |
| Smoking |  |  |  |
|  | Never, n (%) | 9,638 (20.45) | 61,212 (62.72) |
|  | Former, n (%) | 14,359 (30.47) | 14,145 (14.49) |
|  | Current, n (%) | 23,126 (49.08) | 22,232 (22.78) |
| Alcohol drinking |  |  |  |
|  | Never, n (%) | 2,939 (6.24) | 25,691 (26.33) |
|  | Occasionally, n (%) | 37,182 (78.90) | 69,787 (71.51) |
|  | At least once per week, n (%) | 5,735 (12.17) | 1,778 (1.82) |
|  | Daily, n (%) | 1,267 (2.69) | 333 (0.34) |
| Fruit and vegetable intake |  |  |  |
|  | Never, n (%) | 818 (1.74) | 711 (0.73) |
|  | 1 or 2 days per week, n (%) | 10,116 (21.47) | 14,442 (14.80) |
|  | 3 or 4 days per week, n (%) | 14,057 (29.83) | 26,495 (27.15) |
|  | 5 or more days per week, n (%) | 22,132 (46.97) | 55,941 (57.32) |
| Body mass index, kg/m<sup>2</sup>, mean (SD) |  | 27.9 (4.2) | 29.5 (5.2) |
| Leisure time physical activity |  |  |  |
|  | No, n (%) | 33,060 (70.16) | 79,443 (81.41) |
|  | Yes, n (%) | 14,063 (29.84) | 18,146 (18.59) |

SD is standard deviation. IQR is inter-quartile range. N is number.

### HDL cholesterol concentration and all-cause mortality

Figure 1 shows the associations of HDL cholesterol concentration as a continuous variable with all-cause mortality. Men were followed for 18.7 (5.0) years [mean (standard deviation)], and there were 11,264 deaths during 879,005 person-years of follow-up. The association of HDL cholesterol concentration with all-cause mortality in men was J-shaped, with higher concentrations associated with greater risks. The HDL cholesterol concentrations associated with the lowest risks of all-cause mortality in men were within a relatively broad nadir around the mean value of 0.93 mmol/L. Women were followed for 19.3 (4.1) years, and there were 15,468 deaths during 1,878,867 person-years of follow-up. The association of HDL cholesterol concentration with all-cause mortality was U-shaped, with both lower and higher concentrations associated with greater risks. The concentrations associated with the lowest risks of all-cause mortality in women were within a relatively narrow nadir around the mean value of 1.03 mmol/L.

**Figure 1.**
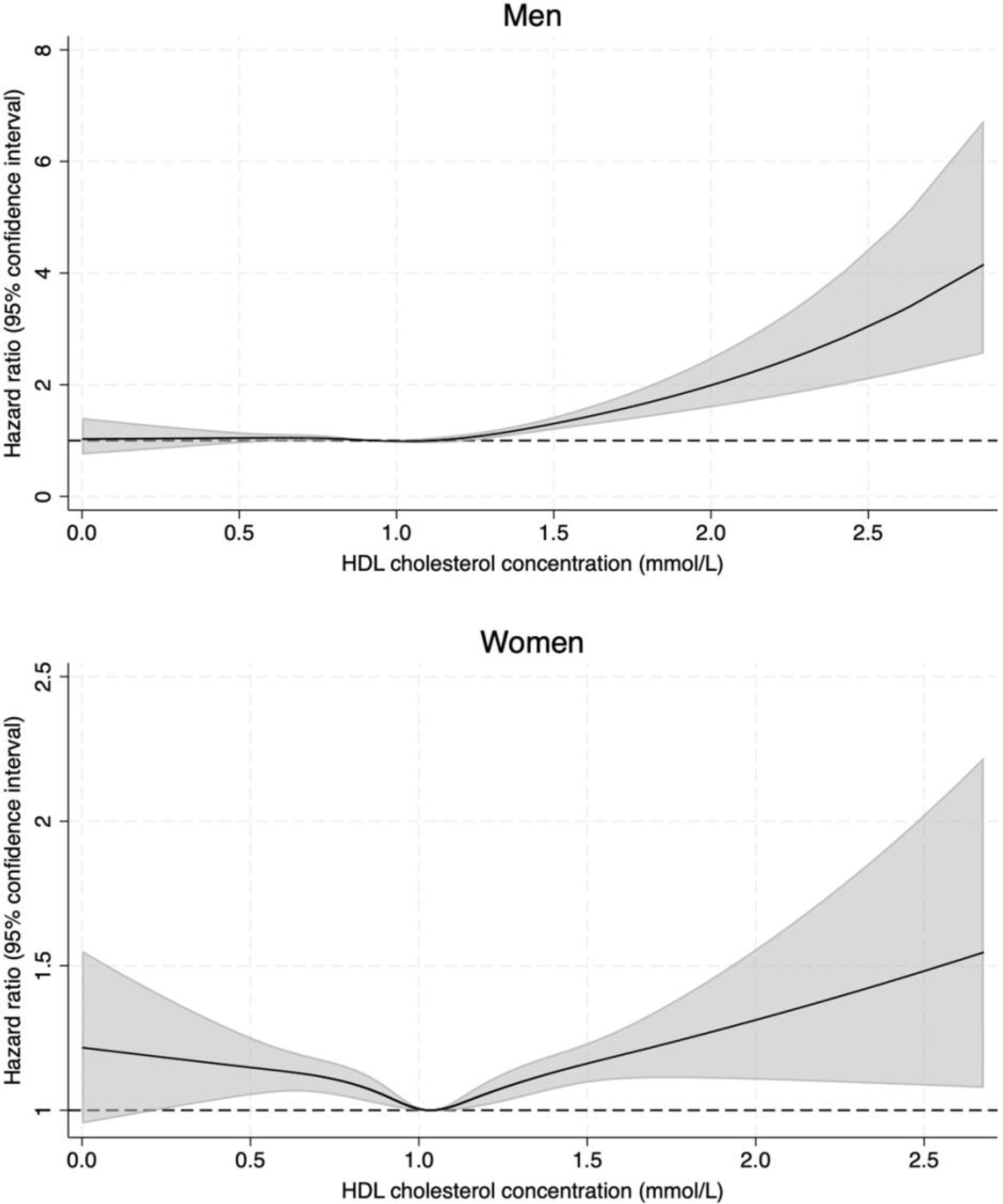
Associations of HDL cholesterol concentration as a continuous variable with all-cause mortality in 47,123 men and 97,589 women. The solid black line is the hazard ratio and the grey shading is the 95% confidence interval. The graphs are from restricted cubic spline regression with three knots, as described in the statistical methods. The graphs are truncated at the 1^st^ and 99^th^ percentiles. Hazard ratios were from Cox regression models adjusted for age, triglyceride concentration, LDL cholesterol concentration, lipid medication, systolic blood pressure, diabetes, education, income, area of residence, smoking, alcohol, fruit and vegetable intake, body mass index, and leisure time physical activity at baseline. Data from the first two years of follow-up were not used to minimise the possibility of reverse causation.

### HDL cholesterol concentration and cause-specific mortality

Figure 2 shows associations of HDL cholesterol concentration as a categorical variable with all-cause mortality, CVD mortality, cancer mortality, and other mortality in men. The reference group was the percentile group containing the HDL cholesterol concentration associated with the lowest risk of all-cause mortality as determined in the restricted cubic spline analysis: the >40^th^-60^th^ percentile group. Similar to the continuous analysis, there was evidence of a J-shaped association of HDL cholesterol concentration with all-cause mortality in men. Compared with the reference group, the multivariate hazard ratio for all-cause mortality (95% confidence interval) was 1.09 (0.99, 1.20) (not significant) in the >95^th^-99^th^ percentile group, 1.54 (1.31, 1.82) (P<0.05) in the >99^th^-100^th^ percentile group, and 1.17 (1.06, 1.28) (P<0.05) in the two highest percentile groups combined. There was no apparent association of HDL cholesterol concentration with CVD mortality after adjusting for potential confounders. Rather, the J-shaped association of HDL cholesterol concentration with all-cause mortality in men was driven by other mortality. Compared with the reference group, the multivariate hazard ratio for other mortality was 1.27 (1.11, 1.44) (P<0.05) in the >95^th^-99^th^ percentile group, 1.82 (1.45, 2.28) (P<0.05) in the >99^th^-100^th^ percentile group, and 1.34 (1.19, 1.52) (P<0.05) in the two highest percentile groups combined. There was also evidence that higher HDL cholesterol concentration was associated with lower risk of cancer mortality. For example, compared with the reference group, the multivariate hazard ratio for cancer mortality was 0.67 (0.49, 0.91) (P<0.05) in the two highest percentile groups combined.

**Figure 2.**
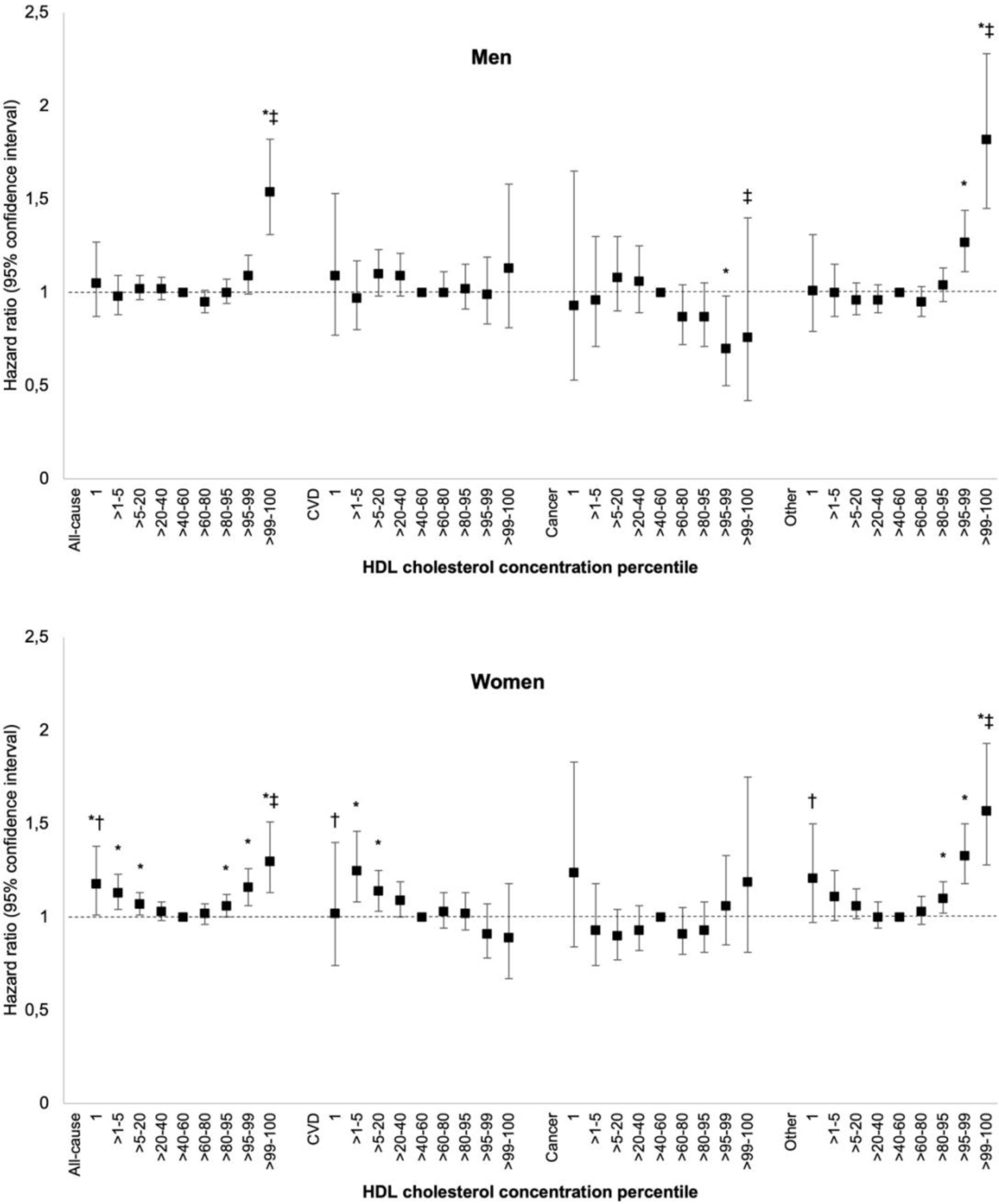
Associations of HDL cholesterol concentration percentiles with mortality in 47,123 men and 97,589 women. CVD deaths, cancer deaths, and other deaths were treated as competing events. All hazard ratios were adjusted for age, triglyceride concentration, LDL cholesterol concentration, lipid medication, systolic blood pressure, diabetes, education, income, area of residence, smoking, alcohol, fruit and vegetable intake, body mass index, and leisure time physical activity at baseline. Data from the first two years of follow-up were not used to minimise the possibility of reverse causation. The reference group was the one containing the HDL cholesterol concentration associated with the lowest risk of all-cause mortality as determined in the restricted cubic spline analysis: >40-60 percentile in both men and women. *Significantly different to reference group. †Average of two lowest percentile groups significantly different to reference group. ‡Average of two highest percentile groups significantly different to reference group. Supplementary Table 1 and Supplementary Table 2 in the online supplement show the cholesterol concentrations in each percentile group, the number of participants, the number of deaths, the incidence rates, and the hazard ratios.

Figure 2 also shows associations of HDL cholesterol concentration as a categorical variable with mortality in women. The reference group was the percentile group containing the concentration associated with the lowest risk of all-cause mortality as determined in the restricted cubic spline analysis: the >40^th^-60^th^ percentile group. Just like the continuous analysis, there was evidence of a U-shaped association of HDL cholesterol concentration with all-cause mortality in women. Indeed, the three lowest percentile groups and the three highest percentile groups were associated with greater risks. The U-shaped association of HDL cholesterol concentration with all-cause mortality in women was largely driven by CVD mortality at lower concentrations and other mortality at higher concentrations. For example, compared with the reference group, the multivariate hazard ratio (95% confidence interval) for CVD mortality was 1.23 (1.05, 1.43) (P<0.05) in the two lowest percentile groups combined; And, the multivariate hazard ratio for other mortality was 1.33 (1.18, 1.50) (P<0.05) in the two highest percentile groups combined. There was no statistically significant association of HDL cholesterol concentration with cancer mortality after adjusting for potential confounders.

### HDL particle count and cause-specific mortality

Table 2 shows associations of small, medium, and large HDL particle counts with mortality in men. Higher small HDL particle count was associated with reduced risk of all-cause mortality, CVD mortality, and other mortality. For example, compared with the bottom quartile of particle count, the multivariate hazard ratio (95% confidence interval) for all-cause mortality was 0.79 (0.73, 0.86) (P<0.05) in the top quartile. Conversely, higher large HDL particle count was associated with increased risk of all-cause mortality and other mortality. For example, compared with the bottom quartile, the multivariate hazard ratio for all-cause mortality was 1.18 (1.12, 1.25) (P<0.05) in the top quartile. Both higher medium HDL particle count and higher large HDL particle count were associated with reduced risk of cancer mortality.

**Table 2.**
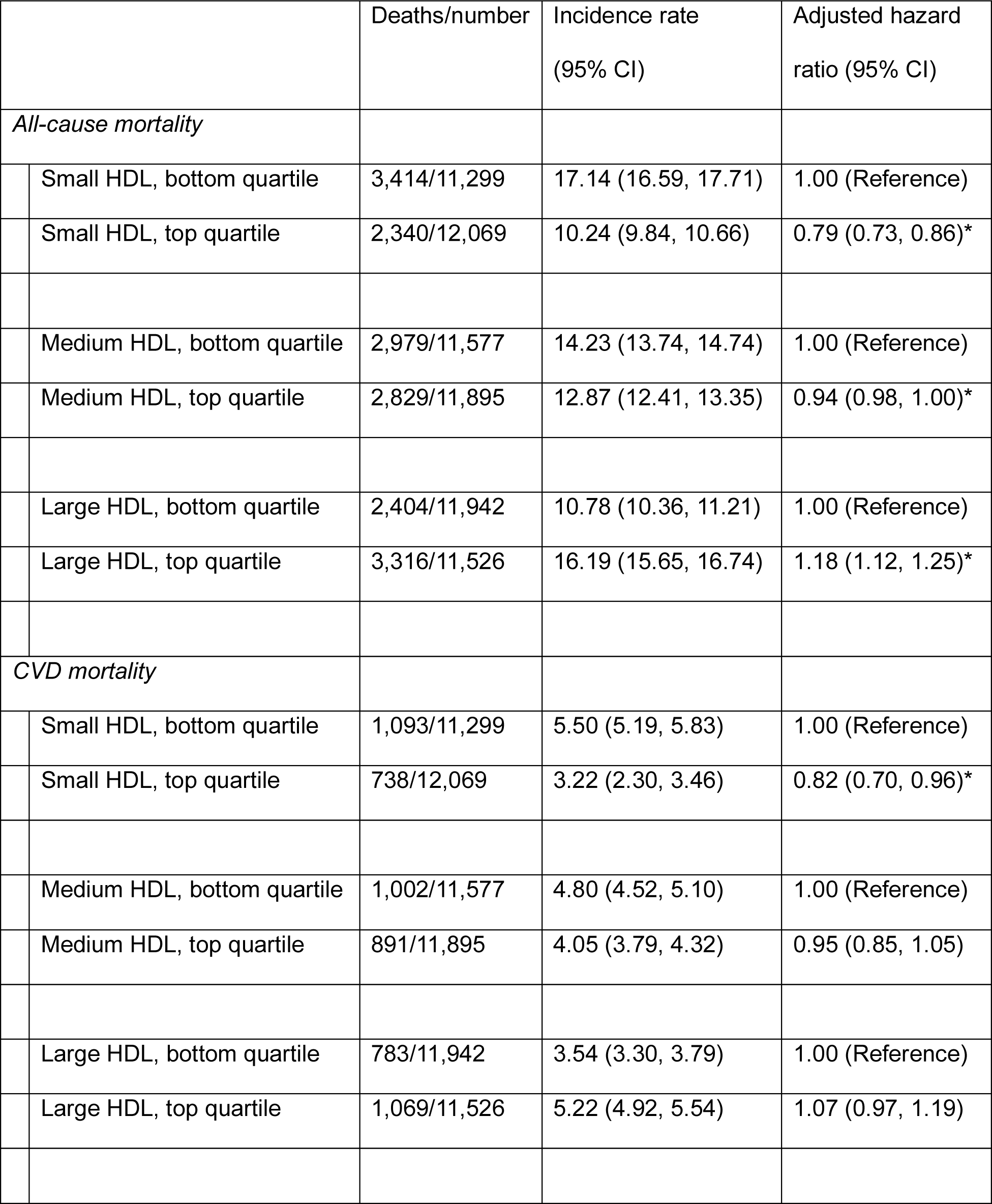
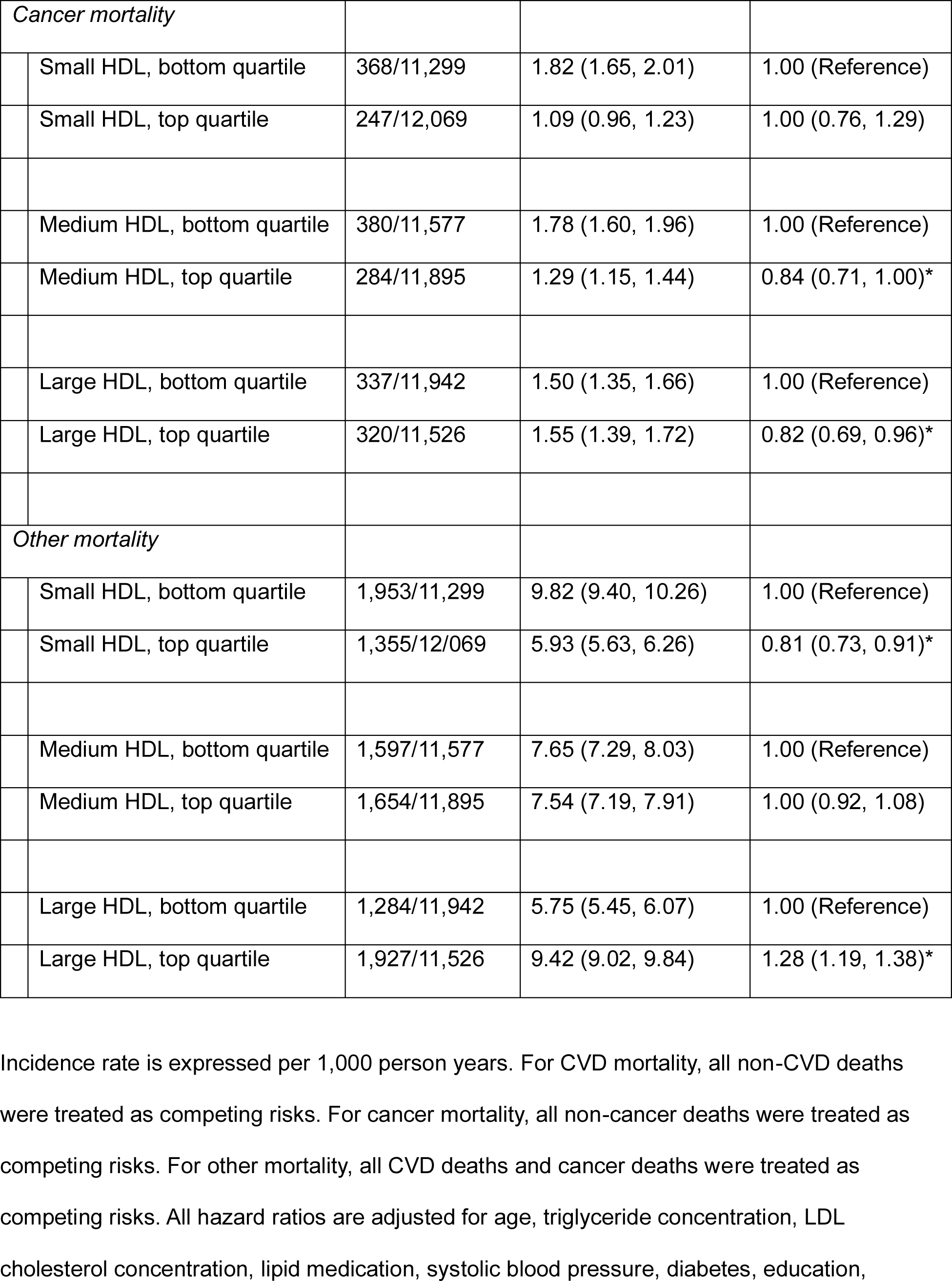
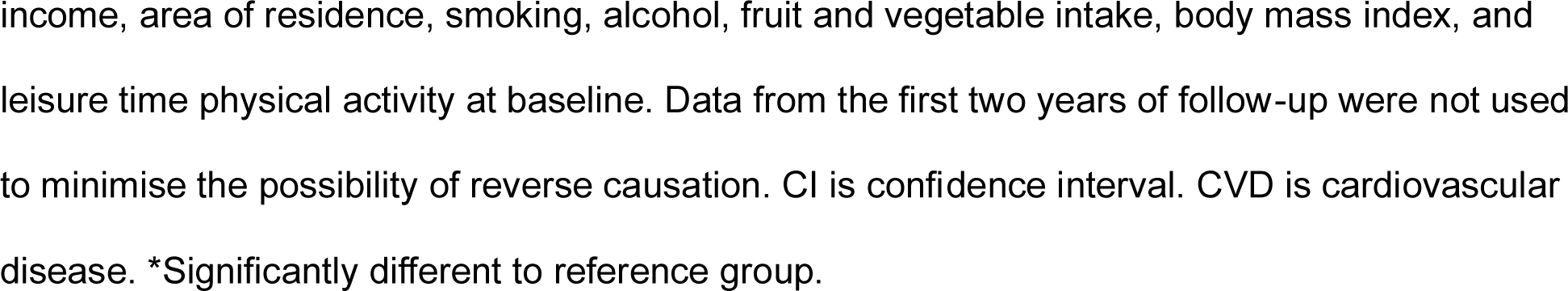
Associations of small, medium, and large HDL particle counts with mortality in men. Incidence rate is expressed per 1,000 person years. For CVD mortality, all non-CVD deaths were treated as competing risks. For cancer mortality, all non-cancer deaths were treated as competing risks. For other mortality, all CVD deaths and cancer deaths were treated as competing risks. All hazard ratios are adjusted for age, triglyceride concentration, LDL cholesterol concentration, lipid medication, systolic blood pressure, diabetes, education, income, area of residence, smoking, alcohol, fruit and vegetable intake, body mass index, and leisure time physical activity at baseline. Data from the first two years of follow-up were not used to minimise the possibility of reverse causation. CI is confidence interval. CVD is cardiovascular disease. *Significantly different to reference group.

Table 3 shows associations of HDL particle counts with mortality in women. Similar to men, higher small HDL particle count was associated with reduced risk of all-cause mortality, CVD mortality, and other mortality. For example, compared with the bottom quartile of particle count, the multivariate hazard ratio (95% confidence interval) for all-cause mortality was 0.77 (0.72, 0.82) (P<0.05) in the top quartile. Also similar to men, higher large HDL particle count was associated with increased risk of all-cause mortality and other mortality. For example, compared with the bottom quartile, the multivariate hazard ratio for all-cause mortality was 1.18 (1.12, 1.23) (P<0.05) in the top quartile. Both higher small particle count and higher medium particle count were associated with increased risk of cancer mortality.

**Table 3.**
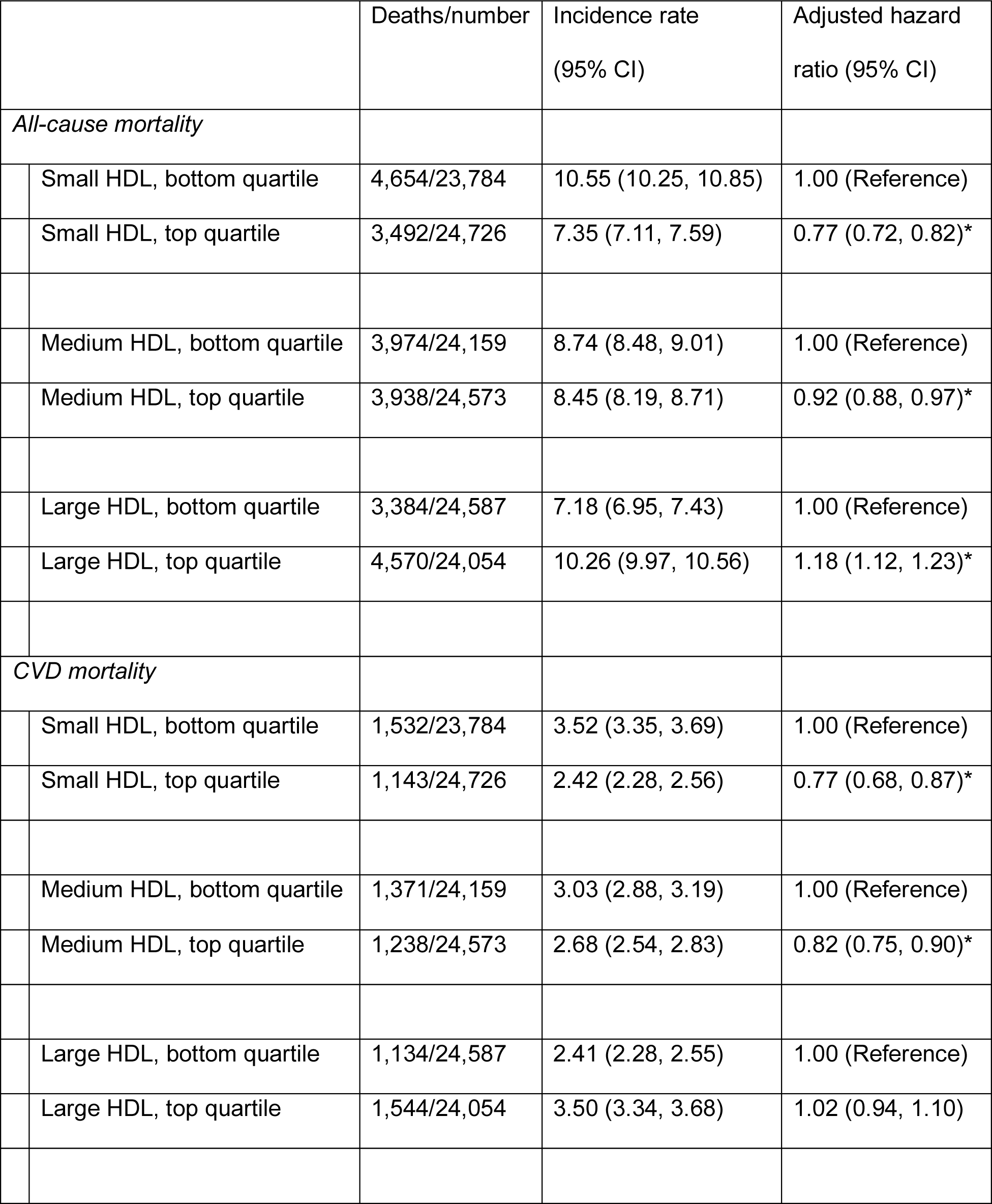
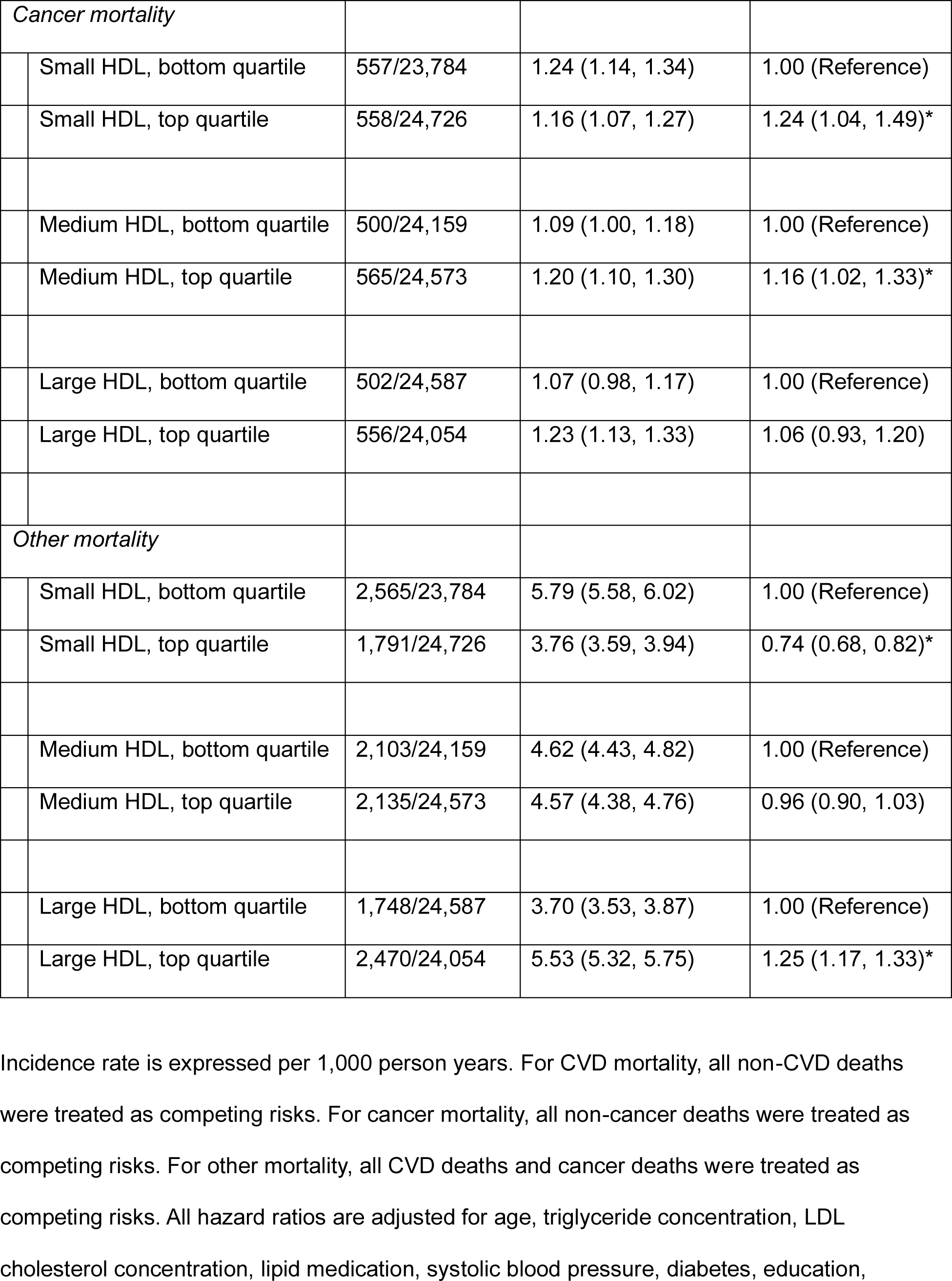
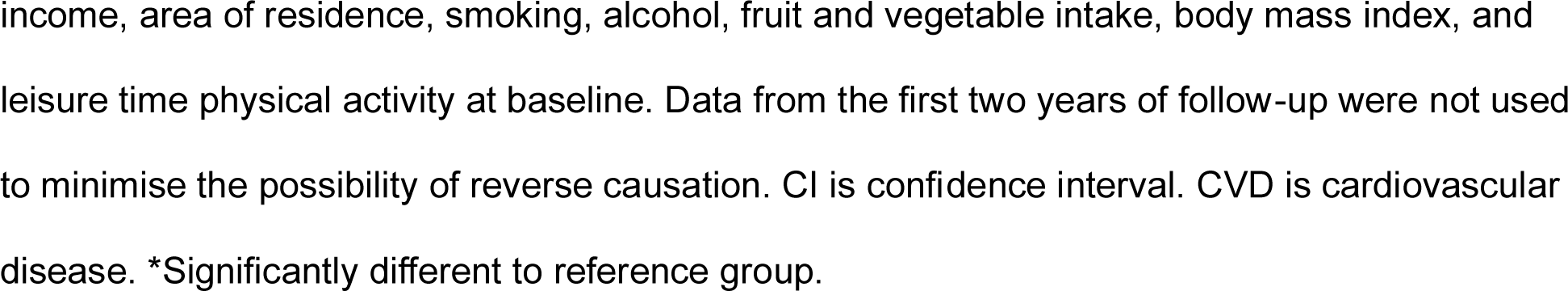
Associations of small, medium, and large HDL particle counts with mortality in women. Incidence rate is expressed per 1,000 person years. For CVD mortality, all non-CVD deaths were treated as competing risks. For cancer mortality, all non-cancer deaths were treated as competing risks. For other mortality, all CVD deaths and cancer deaths were treated as competing risks. All hazard ratios are adjusted for age, triglyceride concentration, LDL cholesterol concentration, lipid medication, systolic blood pressure, diabetes, education,income, area of residence, smoking, alcohol, fruit and vegetable intake, body mass index, and leisure time physical activity at baseline. Data from the first two years of follow-up were not used to minimise the possibility of reverse causation. CI is confidence interval. CVD is cardiovascular disease. *Significantly different to reference group.

## Discussion

The first objective of the present study was to investigate whether there were non-linear associations of HDL cholesterol concentration with all-cause mortality. We found that there was a J-shaped association in men and a U-shaped association in women. The second objective was to investigate what was driving these associations. We found that the J-shaped association of HDL cholesterol concentration with all-cause mortality in men was driven by other mortality; And, we found that the U-shaped association of HDL cholesterol concentration with all-cause mortality in women was largely driven by CVD mortality at lower concentrations and other mortality at higher concentrations. The third objective was to investigate associations of HDL particle counts with mortality. We found that small HDL particle count was associated with reduced risk of all-cause mortality, CVD mortality, and other mortality in both men and women. Conversely, we found that large HDL particle count was associated with increased risk of all-cause mortality and other mortality in men and women. To the best of our knowledge, this is the largest study of the associations of both HDL cholesterol concentration and HDL particle count with mortality.

It is plausible that both low and high HDL cholesterol concentrations are associated with increased risk of mortality [5, 7]. Low concentrations may be associated with increased risk because of a reduction in the beneficial effects of the particle. HDL cholesterol has many beneficial effects that are exerted through three major mechanisms. First, HDL cholesterol has beneficial effects on atherosclerosis that are exerted through reverse cholesterol transport where the particle brings about cholesterol efflux from the macrophage foam cells of atherosclerotic plaques [7]. Second, HDL cholesterol has anti-oxidant properties and anti-inflammatory properties that are exerted through the particle’s many roles in communication with cells [7]. Third, HDL cholesterol has beneficial effects on the immune system that are likely exerted through the particle’s ability to bind to and inactivate potentially toxic substances like bacterial lipopolysaccharides and oxidized lipids [7]. High HDL cholesterol concentrations may be associated with increased risk of mortality because small functional particles may become large dysfunctional particles [7]. When the particle becomes dysfunctional, there is a reduction in reverse cholesterol transport, an increase in inflammatory activity, and an increase in oxidized lipids [7]. When HDL cholesterol concentrations are very high, it may result in endothelial dysfunction because of irregular apoptosis, a reduction in nitric oxide production, or an increase in vascular cell adhesion molecule expression in endothelial cells [7]. It should be stressed that the measures of HDL cholesterol concentration typically used in large observational studies do not reflect the beneficial properties or functions of the particle [7].

The present study may help increase our understanding of the associations of HDL cholesterol with mortality because it includes measure of both particle concentration and particle count. It is interesting that there was a broad nadir and a J-shaped association of HDL cholesterol concentration with all-cause mortality in men because it would suggest that changes in concentration do not lead to changes in risk in the vast majority of men in Mexico City; Rather, increases in concentration only lead to increases in risk in the minority of men on the right end of the J-shaped curve. The fact that other mortality drove the J-shaped association would suggest that high HDL cholesterol concentration plays a role in non-CVD and non-cancer deaths in men. HDL particle size is larger at higher HDL cholesterol concentrations [16] and it has been suggested that large HDL particles may interfere with the actions of innate and adaptive immune cells and increase the risk of autoimmune and infectious diseases [7]. The present study suggests that large HDL particle count is indeed associated with increased risk of other mortality in men. It is also interesting that there is a narrow nadir and a U-shaped association in women because it would suggest that the vast majority of women in Mexico City are susceptible to changes in HDL cholesterol concentrations; Indeed, decreases in concentration lead to increases in risk along the left of the U-shaped curve and increases in concentration lead to increases in risk along the right of the curve. The fact that CVD mortality drove the U-shaped association at lower concentrations would suggest that low HDL cholesterol concentration plays a role in atherosclerotic cardiovascular disease in women. In fact, low HDL cholesterol concentration is strongly associated with increased levels of the triglyceride-rich lipoproteins [37] that impair cholesterol metabolism [38]. Given that other mortality drove the U-shaped association at higher concentrations, it would appear that high HDL cholesterol concentrations increase the risk of autoimmune and infectious diseases in both men and women. Genetic variation may also help explain the associations observed in men and women in the present study because 19% of adults with low HDL cholesterol concentrations and 11% of adults with high concentrations may have variants in HDL genes [39] that could impair HDL function and, theoretically, increase the risk of death [5].

The available evidence has important implications for policy and practice because higher HDL cholesterol concentrations are still regarded as desirable in Mexico [40] and other countries [1–3]. The first studies to find that there were U-shaped associations of HDL cholesterol concentration with all-cause mortality were conducted in Europe [10, 15], and there were immediate calls for more research in other regions [15, 41]. Since then, original studies in North America [11, 12, 14] and Western Pacific [9, 12], and a pooled analysis of studies in Europe, North America, and Western Pacific [42], have shown that there are U-shaped or J-shaped associations of HDL cholesterol concentration with mortality. Consistency of association is an important consideration in epidemiology [43], and to the best of our knowledge, this is the first study in Latin America to find that there are U-shaped or J-shaped associations of HDL cholesterol concentration with mortality. Regardless of whether the associations are U-shaped or J-shaped and regardless of whether CVD deaths, cancer deaths, or other deaths drive the associations, it would now seem that there is little justification for the raising of HDL cholesterol concentration as a therapeutic target because higher concentrations are associated with increased risk of death. While the raising of HDL cholesterol concentration may not be an appropriate therapeutic target, the present study suggests that HDL particle count may be an important biomarker. Indeed, we found that small HDL particle count was associated with reduced risk of all-cause mortality, CVD mortality, and other mortality in both men and women. Conversely, we found that large HDL particle count was associated with increased risk of all-cause mortality and other mortality in men and women. A recent study of 103,265 adults in Europe also found that higher small HDL particle count was associated with reduced risk of all-cause mortality [16]. Another study of 30,195 adults in Europe would suggest that the associations of small HDL particle count with mortality are partly driven by infectious disease deaths: higher small-and-medium HDL particle count was associated with lower risk of infectious disease-related death after adjusting for age, sex, lipid medication, and other potential confounders [44].

This study has some limitations. The Mexico City Prospective Study was deemed to be representative of the male and female populations aged 35 years or older because of the large sample sizes and the high response rates, but there were no formal tests of representativeness [17]. Participants were not asked to fast; however, the non-fasting state may be preferable in epidemiological studies because it is more indicative of the normal lipid profile [45]. Furthermore, HDL cholesterol concentration was associated with mortality in another large study in which the same NMR platform was used and the average time since the last meal was also around four hours [19]. Some variables were self-reported, which may introduce bias. The assessment of physical activity was relatively crude, but simple questionnaires are sufficient to identify the physically active [46]. The assessment of diet was also relatively crude, but fruit and vegetable intake is associated with overall diet quality [47]. It is unclear why HDL cholesterol concentration and large HDL particle count were associated with reduced risk of cancer mortality in men in the present study; however, the results should be treated with caution because there were relatively few cancer deaths in men. Prospective studies have produced mixed results [48], and more research has been called for to clarify the role of HDL in cancer risk [5]. HDL’s are heterogenous particles with many biological functions [7, 49], and more research is required to understand the associations of particle size with mortality and to determine whether particle function can be maintained or improved with lifestyle interventions or pharmacological interventions [6].

In conclusion, this is the largest study of the associations of both HDL cholesterol concentration and HDL particle count with mortality. The J-shaped and U-shaped associations observed in this study in Latin America and other studies in different regions suggest that raising HDL cholesterol concentration is not an appropriate therapeutic target. More research is required to confirm the novel finding that small HDL particle count is associated with reduced risk of all-cause mortality, CVD mortality, and other mortality in both men and women. More research is also required to confirm the novel finding that large HDL particle count is associated with increased risk of all-cause mortality and other mortality in men and women.

## Data Availability

Data from the Mexico City Prospective Study are available to bona fide researchers. The study's Data and Sample Sharing policy can be downloaded (in English or Spanish): https://www.ctsu.ox.ac.uk/research/mexico-city-prospective-study. Available study data can be examined in detail using the study's Data Showcase: https://datashare.ndph.ox.ac.uk/mexico/. Ancestry-specific allele frequencies are also available in a public browser: https://rgc-mcps.regeneron.com/license-and-terms-of-use.

https://datashare.ndph.ox.ac.uk/mexico/

https://rgc-mcps.regeneron.com/license-and-terms-of-use

## Acknowledgements

The authors thank everyone who took part in the study. The data used in this analysis were obtained through an open-access data request made to the Mexico City Prospective Study (MCPS) principal investigators. This analysis was conducted under MCPS Data Application Numbers 2023-029 and 2024-009.

